# Association of circulating endothelial progenitor cell count and functional outcome in patients with acute ischemic stroke due to intracranial large vessel occlusion

**DOI:** 10.64898/2026.06.11.26355469

**Authors:** Ana Aguilera-Simón, Pol Camps-Renom, Marina Guasch-Jiménez, Núria Puig, Elena Jiménez-Xarrié, Rebeca Marín, Marta Soler, Cristina Gallego-Fabrega, Garbiñe Ezcurra-Díaz, Álvaro Lambea-Gil, Alejandro Martínez-Domeño, Luis Prats-Sánchez, Anna Ramos-Pachón, José Pablo Martínez-González, Joaquín Ortega-Quintanilla, Joan Martí-Fàbregas

## Abstract

**Background:** Circulating endothelial progenitor cells (cEPCs) contribute to vascular repair following an ischemic stroke. The aim of the study was to evaluate the association between cEPCs and functional outcomes in patients with acute ischemic stroke (AIS) due to large vessel occlusion (LVO) who received endovascular therapy (EVT).

**Methods:** Prospective study of patients with LVO-AIS who received EVT. Blood samples were obtained within 24±12 hours and on day 7±1 from stroke onset. cEPCs were detected using flow cytometry (CD34+/VEGFR2+/CD133+). The primary endpoint was a favourable functional outcome (modified Rankin Scale 0-2) at three months’ follow-up. Secondary endpoints include baseline to 24 hours/day 7 changes in the National Institutes of Health Stroke Scale (NIHSS) score and collateral circulation (CC) status. Bivariate and multivariable logistic regression analyses were performed.

**Results:** Included were 90 patients (73.2±12.7 years, 41.1% women) in 42 of whom (46.7%) cEPCs were detected at 24 hours. On day 7, cEPCs were detected in 27 (43.6%) of 62 patients for which this information was available. Atrial fibrillation, prior anticoagulant treatment and stroke onset-to-door time <6 hours were associated with lower cEPC counts, and intravenous fibrinolysis therapy was associated with a higher cEPC count on day 7. No association was found between cEPCs and functional outcomes at three months. Patients with the highest cEPC count (Q4) at 24 hours had a lower probability of good CC (46.2% vs 77.3%; p=0.031).

**Conclusion:** cEPC count in patients with LVO-AIS who received EVT was not associated with functional outcomes.

## Introduction

Circulating endothelial progenitor cells (cEPCs) are stem cells derived from bone marrow that circulate in the peripheral blood. After an ischemic stimulus, cEPCs migrate from the bone marrow to the damaged tissue where, differentiated into mature endothelial cells, they contribute to vascular repair and the formation of new blood vessels[1–3]. Vascular risk factors, such as age and hypertension, may reduce cEPC mobilization[4], whereas treatment with statins[5], antiplatelets[6], and antihypertensives[7,8] can enhance cEPC mobilization.

Although findings are controversial, several studies have reported that increased cEPC levels are associated with low cardiovascular risk[9–11] and better clinical outcomes for mild ischemic stroke[12]. However, this association has not been yet studied for acute ischemic stroke (AIS) due to large vessel occlusion (LVO).

We hypothesize that cEPC mobilization may be associated with functional outcomes following LVO-AIS through two possible mechanisms. One mechanism is that cEPCs, due to their role in vascular homeostasis, may improve clinical outcomes through the release of angiogenic[13], vasodilatory[14], and anti-inflammatory[15] factors, reducing infarct growth[12] and mediating in endothelial cell regeneration and neovascularization[1],[2]. The other mechanism is the critical role played by cerebral collateral circulation (CC) during the brain ischemia acute phase [16], as better CC has been associated with better clinical outcomes in patients receiving endovascular therapy (EVT)[17]. To date, however, little is known about factors influencing cerebral CC and, accordingly, we hypothesized that cEPCs could also be related to the degree and maintenance of cerebral CC in patients with LVO-AIS.

The aim of this study was to investigate whether early cEPC mobilization was associated with clinical outcomes in patients with AIS secondary to LVO who received EVT and whether this association was mediated by cerebral CC status.

## Methods

### Study design

A prospective study of consecutive patients with AIS due to anterior circulation LVO who underwent EVT was carried out in our Comprehensive Stroke Centre between May 2020 and May 2023. The study was approved by the Ethics Committee of the Hospital de la Santa Creu i Sant Pau and the patients, or their legal representative, gave their written informed consent to participate. The study was performed in accordance with the Declaration of Helsinki.

### Study population

The inclusion criteria were as follows: (1) age ≥18 years; (2) anterior circulation AIS; (3) LVO of the M1 or M2 segments of the middle cerebral artery (MCA) or terminal internal carotid artery (TICA), with or without significant stenosis or occlusion of the extracranial internal carotid artery; (4) EVT performed within the first 24 hours after stroke onset or the last time seen well; (5) previous modified Rankin Scale (mRS) score 0-2. Exclusion criteria were occlusion of other arterial segments (vertebrobasilar circulation, A1, P1, M3, M4).

The following variables were recorded for all patients: demographic data (age and sex); vascular risk factors (previous stroke, arterial hypertension, diabetes, dyslipidemia, smoking habits, obesity, ischemic heart disease, heart failure, atrial fibrillation); drug treatment on admission (antiplatelets, anticoagulants, statins); previous mRS score; National Institutes of Health Stroke Scale (NIHSS) score on admission; etiology according to the Trial of Org 10172 (TOAST) criteria; Alberta Stroke Program Early CT Score (ASPECTS); intravenous thrombolysis therapy; modified Thrombolysis in Cerebral Infarction (mTICI) score at procedure end; and stroke onset-to-groin puncture time. Tandem lesions were classified according to occlusion of the intracranial artery (M1 or M2 segments of the MCA, or TICA).

### cEPC measurement

Blood samples, collected in ethylene diamine tetra acetic acid (EDTA) tubes, were obtained within 24±12 hours and 7±1 days from stroke onset. Erythrocytes were first lysed using a red blood lysis solution. cEPCs were then detected through specific surface antigens (CD34+/VEGFR2+/CD133+) using magnetic-activated cell sorting (MACS). The method was then refined using a magnetic pre-enrichment system (EPC Enrichment and Enumeration Kit-Human, Miltenyi Biotec, Germany). Briefly, cells were magnetically labelled before fluorescent staining and then, using a MACSQuant flow cytometer (Miltenyi Biotec, Germany), were retained in a magnetic column before being eluted and counted. Finally, having settled on the appropriate gate for mononuclear cells, triple-positively-stained cells were identified. Results were expressed as cEPCs as a proportion of the total number of mononuclear cells. As previously described in the literature, an absolute change of ≥0.01% in the cEPC count was considered to be a significant variation (increase or decrease)[18].

### Imaging analysis

All patients underwent brain computed tomography (CT) and CT-angiography (CTA) on admission. A certified neuroradiologist blinded to the outcomes and cEPC results analysed the images and generated a collateral score (CS) for CC assessment, ranging from 0 to 3 according to the Tan et al. [19] scale, which compares arterial filling of affected and unaffected hemispheres. CS was thus graded according to occluded territory filling as follows: 0, no filling; 1, collateral filling ≤50%; 2, collateral filling >50% but <100%; and 3, collateral filling 100%[19]. Dichotomizing the CS scores, we classified 0 or 1 as poor CC, and 2 or 3 as good CC.

The CS analysis was performed after patient care and exclusively for research purposes; it was not used to select patients for EVT.

### Primary and secondary endpoints

The primary endpoint was functional outcome at three months’ follow-up assessed by mRS scoring, dichotomized as favourable (mRS 0-2) or unfavourable (mRS 3-6). Secondary outcomes were change from baseline to 24 hours in the NIHSS score (ΔNIHSS_b–24h_), change from baseline to day 7 in the NIHSS score (ΔNIHSS_b–d7_), and CS score for the baseline CTA.

### Statistical analysis

Continuous variables were reported as means and standard deviations (SD), or as medians and interquartile range (IQR) if non-normally distributed, as tested by the Shapiro-Wilk normality test. Categorical variables were expressed as counts and percentages. As cEPC counts were not normally distributed and a high percentage of patients had no detectable cEPCs, the study population was divided into cEPC presence (cEPCs+) and absence (cEPCs-) and by cEPC quartiles (Q1-Q4).

We first described the clinical characteristics of the population and studied associations between cEPC count and clinical variables. Using the Mann-Whitney U test for continuous variables and the chi-square or Fisher exact test for categorical variables, we conducted bivariate analyses comparing the following: cEPC presence/absence at 24 hours, cEPC presence/absence on day 7, and change in the cEPC count in the first week after stroke onset (ΔcEPCs_24h–d7_).

We next compared cEPC counts at different times (cEPCs +/-24 hours, cEPCs day 7, and ΔcEPCs +/-) with functional outcomes at three months’ follow-up. Also, since cEPC mobilization is known to be time-dependent, we performed a subanalysis of patients whose cEPC counts were obtained strictly within the first 24 hours after stroke onset (cEPCs <24 hours). For this purpose we performed bivariate analyses, using the Mann-Whitney U test for continuous variables and the chi-square test for categorical variables.

Finally, we tested associations between cEPC counts and good/poor CC, performing bivariate analyses of predictors (including cEPC count) using the Student’s t-test for continuous variables (or the Mann-Whitney U test for non-parametric variables) and the chi-square test or Fisher exact test for categorical variables. To test the variables independently associated with the probability of good/poor CC, we implemented multivariable logistic regression analysis using a backward stepwise selection approach, dichotomizing cEPCs in terms of absence/presence and by quartile.

The results were expressed as odds ratio (OR) and 95% confidence interval (CI) values. Statistical significance for all the analyses was set to p<0.05 (two-sided). Analyses were performed using Stata v.17 (Texas, USA).

## Results

### Clinical characteristics

A total of 90 patients were included, mean age 73.2±12.7 years, 37 (41.1%) women. Previous mRS median score was 0 (IQR 0-1) and the baseline NIHSS median score was 17 (IQR 11-21). Clinical characteristics of the population are detailed in Table 1.

**Table 1.** Clinical characteristics.

|  | **n=90** |
| --- | --- |
| Age, mean (SD) | 73.2 (12.7) |
| Sex (women), n (%) | 37 (41.1) |
| Current smokers, n (%) | 17 (18.9) |
| Obesity (BMI $\geq 30$), n (%) | 16 (17.8) |
| Hypertension, n (%) | 62 (68.9) |
| Diabetes, n (%) | 21 (23.3) |
| Dyslipidemia, n (%) | 48 (53.3) |
| Previous stroke, n (%) | 16 (17.8) |
| Heart failure, n (%) | 11 (12.2) |
| Atrial fibrillation, n (%) | 24 (26.7) |
| Prior antiplatelet treatment, n (%) | 24 (26.7) |
| Prior statin treatment, n (%) | 39 (43.3) |
| Prior anticoagulant treatment, n (%) | 24 (26.7) |
| Prior antihypertensive treatment, n (%) | 59 (65.6) |
| Previous mRS score, median (IQR) | 0 (0-1) |
| Baseline NIHSS, median (IQR) | 17 (11-21) |
| ASPECTS, median (IQR)* | 9 (7-10) |
| Stroke onset-to-door time <6 hours, n (%) | 64 (71.1) |
| Stroke onset-to-groin puncture time (minutes), median (IQR) | 196 (109-442) |
| Occlusion, n (%) |  |
| M1 | 48 (53.3) |
| M2 | 26 (28.9) |
| TICA | 16 (17.8) |
| Stroke etiology, n (%)* |  |
| Large-artery atherosclerosis | 16 (18.0) |
| Cardioembolism | 41 (46.1) |
| Uncommon | 7 (7.9) |
| Undetermined | 25 (28.1) |
| Reperfusion therapy, n (%) |  |
| Mechanical thrombectomy | 59 (65.6) |
| Intravenous fibrinolysis and mechanical thrombectomy | 31 (34.4) |
| mTICI score $\geq 2b$, n (%)** | 85 (96.6) |
| Good CC (2-3), n (%)*** | 65 (76.5) |

\*, \*\*, \*\*\*: data available for 89, 88, and 85 patients, respectively. ASPECTS=Alberta Stroke Program Early CT Score; BMI=body mass index; CC=collateral score; IQR=interquartile range; mRS=modified Rankin scale; mTICI=modified Thrombolysis in Cerebral Infarction; NIHSS=National Institute of Health Stroke Scale; SD=standard deviation; TICA=terminal internal carotid artery.

At 24 hours, median cEPCs as a proportion of mononuclear cells was 0% (IQR 0-0.02) and cEPCs were detectable in 42 patients (46.7%). cEPCs were found in 27 (43.6%) of the 62 patients for whom a control blood sample was obtained on day 7, when median cEPCs as a proportion of mononuclear cells was 0% (IQR 0-0.02) and cEPC counts had increased in 15 patients (24.2%). Supplementary Tables S1, S2, and S3 show patient baseline characteristics associated with the probability of cEPCs within 24 hours (cEPCs+ 24 hours) and on day 7 (cEPCs+ day 7), and with the probability of an increased cEPC count in the first week after stroke onset (ΔcEPCs+), respectively.

Remarkably, in the bivariate analysis the only variable associated with cEPC count at 24 hours was previous atrial fibrillation. However, associated with cEPCs count on day 7, in addition to atrial fibrillation, were prior use of anticoagulant treatment, stroke onset-to-door time <6 hours, and intravenous fibrinolysis therapy. Finally, the variables associated with ΔcEPCs in the first week after stroke onset were stroke onset-to-door time <6 hours and intravenous fibrinolysis therapy.

### Primary endpoint

Bivariate analysis results are shown in Table 2. At three months, we had data for 77 patients. The median mRS score was 3 (IQR 1-6) and outcomes were favourable (mRS 0-2) for 36 patients (46.7%).

**Table 2.**
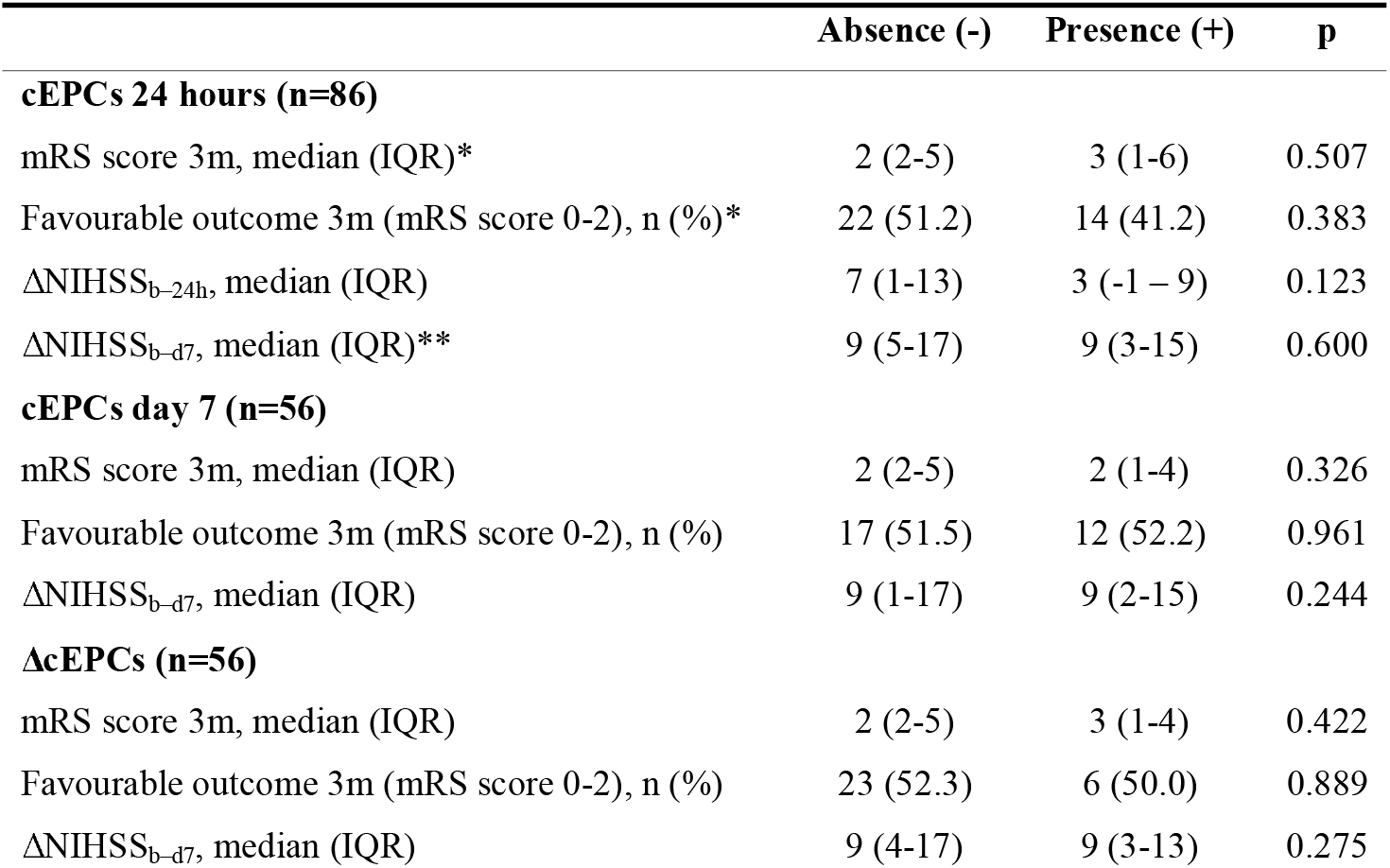
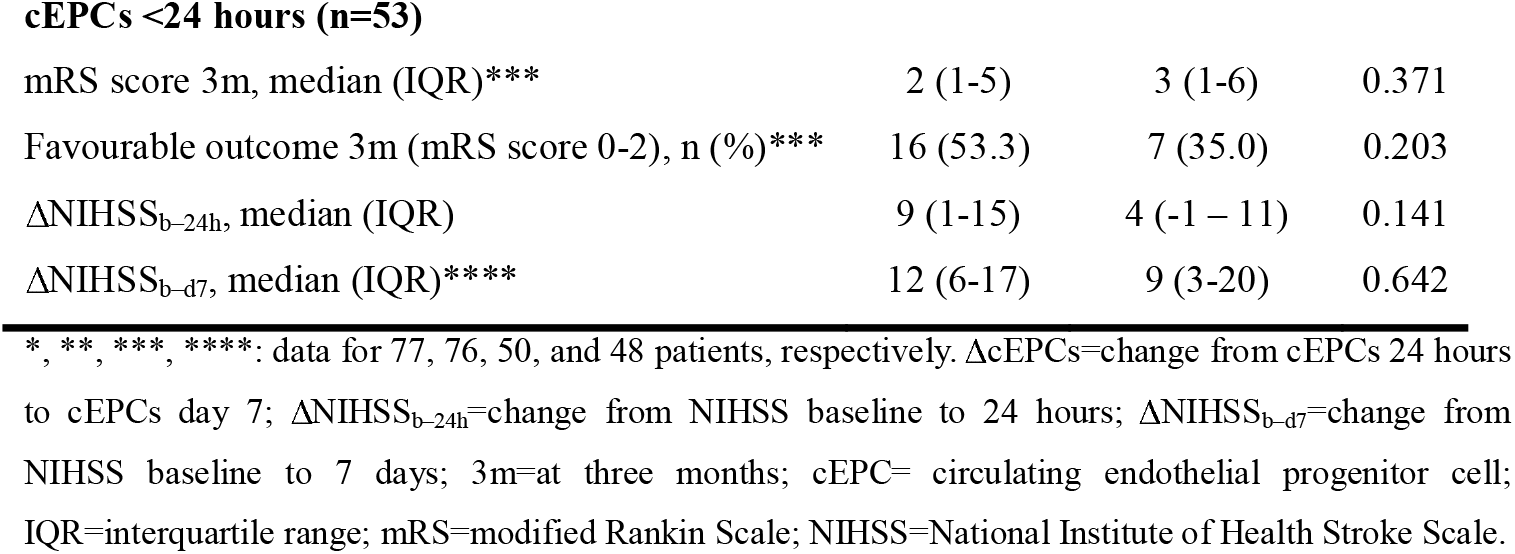
Bivariate analysis between cEPC presence/absence and variables related to functional outcomes at three months.

|  | Absence (-) | Presence (+) | p |
| --- | --- | --- | --- |
| **cEPCs 24 hours (n=86)** |  |  |  |
| mRS score 3m, median (IQR)* | 2 (2-5) | 3 (1-6) | 0.507 |
| Favourable outcome 3m (mRS score 0-2), n (%)* | 22 (51.2) | 14 (41.2) | 0.383 |
| ΔNIHSS<sub>b-24h</sub>, median (IQR) | 7 (1-13) | 3 (-1 – 9) | 0.123 |
| ΔNIHSS<sub>b-d7</sub>, median (IQR)** | 9 (5-17) | 9 (3-15) | 0.600 |
| **cEPCs day 7 (n=56)** |  |  |  |
| mRS score 3m, median (IQR) | 2 (2-5) | 2 (1-4) | 0.326 |
| Favourable outcome 3m (mRS score 0-2), n (%) | 17 (51.5) | 12 (52.2) | 0.961 |
| ΔNIHSS<sub>b-d7</sub>, median (IQR) | 9 (1-17) | 9 (2-15) | 0.244 |
| **ΔcEPCs (n=56)** |  |  |  |
| mRS score 3m, median (IQR) | 2 (2-5) | 3 (1-4) | 0.422 |
| Favourable outcome 3m (mRS score 0-2), n (%) | 23 (52.3) | 6 (50.0) | 0.889 |
| ΔNIHSS<sub>b-d7</sub>, median (IQR) | 9 (4-17) | 9 (3-13) | 0.275 |
| cEPCs <24 hours (n=53) |  |  |  |
| mRS score 3m, median (IQR)*** | 2 (1-5) | 3 (1-6) | 0.371 |
| Favourable outcome 3m (mRS score 0-2), n (%)*** | 16 (53.3) | 7 (35.0) | 0.203 |
| ΔNIHSS<sub>b-24h</sub>, median (IQR) | 9 (1-15) | 4 (-1 – 11) | 0.141 |
| ΔNIHSS<sub>b-d7</sub>, median (IQR)**** | 12 (6-17) | 9 (3-20) | 0.642 |

\*, \*\*, \*\*\*, \*\*\*\*: data for 77, 76, 50, and 48 patients, respectively. ΔcEPCs=change from cEPCs 24 hours to cEPCs day 7; ΔNIHSS<sub>b-24h</sub>=change from NIHSS baseline to 24 hours; ΔNIHSS<sub>b-d7</sub>=change from NIHSS baseline to 7 days; 3m=at three months; cEPC= circulating endothelial progenitor cell; IQR=interquartile range; mRS=modified Rankin Scale; NIHSS=National Institute of Health Stroke Scale.

Favourable outcome rates were similar for patients with and without cEPC mobilization at 24 hours (41.2% vs 51.2%; p=0.383). Results remained unchanged after adjusting for well-known functional outcome predictors (baseline NIHSS, time from symptoms onset, ASPECTS, age, and final mTICI). Likewise, no significant differences were observed in favourable outcome rates for patients with and without detectable cEPCs on day 7 (52.2% vs 51.5%; p=0.961), or for patients with ΔcEPCs+ compared to ΔcEPCs-(50.0% vs 52.3%; p=0.889). cEPC presence/absence was not associated with functional outcomes in multivariable logistic regression analyses adjusted for the abovementioned well-known predictors.

### Secondary endpoint

Bivariate analysis results are shown in Table 2. Patients with detectable cEPC counts at 24 hours from stroke onset tended to have more severe deficits as assessed by ΔNIHSS_b–24h_ (median ΔNIHSS_b–24h_ 7 (IQR 1-13) vs median ΔNIHSS_b–24h_ 3 (IQR -1–9); p=0.123), although the difference was not statistically significant. No significant differences were found either between ΔNIHSS_b–d7_ and cEPC count on day 7 or for ΔcEPCs.

Included in the sensitivity subanalysis of patients with cEPCs <24 hours from stroke onset were 57 patients. The trend observed above persisted, i.e., between early cEPCs mobilization and the probability of more severe deficits at 24 hours, although again, the association was not statistically significant (median ΔNIHSS_b–24h_ 9 (IQR 1-15) vs median ΔNIHSS_b–24h_ 4 (IQR -1– 11); p=0.141).

### cEPCs and CC

Figure 1 shows cEPC count distributions for patients with poor compared to good CC. Patients with, compared to without, measurable cEPCs at 24 hours had a higher probability of presenting poor CC at baseline, although the difference was not significant. Repeating the analysis by cEPC quartiles, patients with the highest cEPC counts (Q4) were found to have a lower probability of good CC (46.2% vs 77.3%; p=0.031) (see Supplementary Table S4; see also Supplementary Table S5 for the bivariate analyses of poor and good CC clinical characteristics).

**Figure 1.**
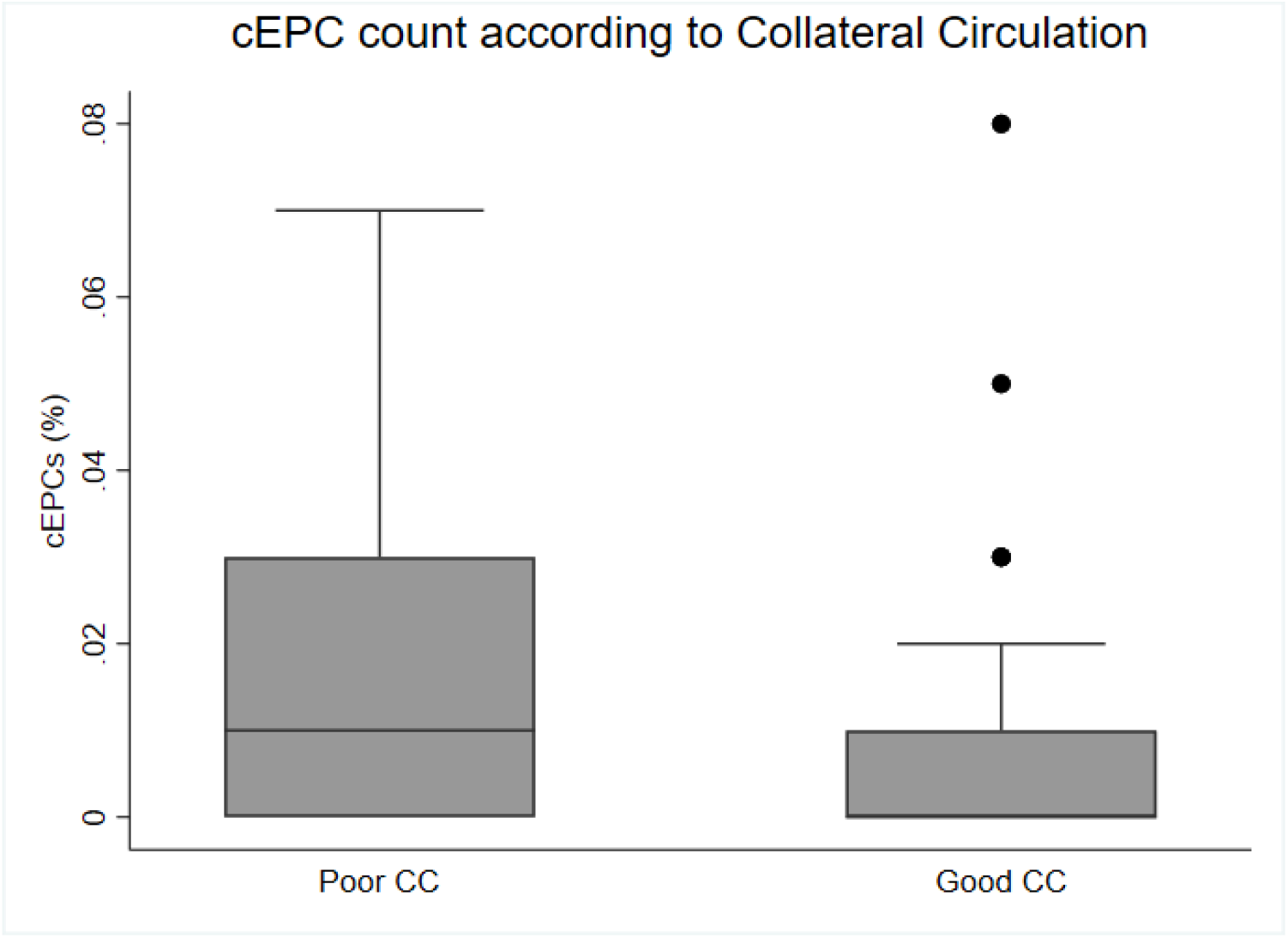
Circulating endothelial progenitor cell (cEPC) count <24 hours and poor/good collateral circulation (CC).

In the multivariable logistic regression analysis, the only variables independently associated with the probability of poor CC were previous atrial fibrillation and greater cEPC mobilization in 24 hours (Q4).

Since treatment with statins, antiplatelets, or antihypertensives is known to potentially modify cEPC mobilization, we performed sensitivity analyses that adjusted for previous treatments. Table 3 reports results adjusted OR (aOR) values for four models. In Model 1, adjusted for prior treatments, higher cEPC counts were associated with a lower probability of good CC (aOR=0.18, 95% CI 0.04-0.82, p=0.027). In Model 2, adjusted additionally for stroke onset-to-groin puncture time, the association persisted between the highest cEPC count (Q4) and a lower probability of good CC (aOR=0.17, 95% CI 0.04-0.80, p=0.025). Similar results were obtained for Model 3 (adjusted additionally for mTICI score) and Model 4 (P adjusted additionally for stroke onset-to-groin puncture time and mTICI score).

**Table 3.** Multivariable logistic regression analysis of predictors of good collateral circulation.

|  | aOR | 95% CI | p |
| --- | --- | --- | --- |
| **Model 1** |  |  |  |
| Q4 | 0.15 | 0.03-0.70 | **0.015** |
| Atrial fibrillation | 0.32 | 0.07-1.38 | 0.127 |
| Prior statin treatment | 0.80 | 0.18-3.49 | 0.763 |
| Prior antiplatelet treatment | 0.50 | 0.12-2.03 | 0.331 |
| Prior antihypertensive treatment | 0.58 | 0.13-2.62 | 0.480 |
| **Model 2** |  |  |  |
| Q4 | 0.15 | 0.03-0.68 | **0.014** |
| Atrial fibrillation | 0.37 | 0.08-1.48 | 0.150 |
| Prior statin treatment | 0.77 | 0.18-3.40 | 0.735 |
| Prior antiplatelet treatment | 0.49 | 0.12-1.98 | 0.318 |
| Prior antihypertensive treatment | 0.55 | 0.12-2.5 | 0.440 |
| Stroke onset-to-groin puncture time | 1.00 | 1.00-1.01 | 0.649 |
| **Model 3** |  |  |  |
| Q4 | 0.12 | 0.03-0.61 | **0.010** |
| Atrial fibrillation | 0.23 | 0.05-1.14 | 0.073 |
| Prior statin treatment | 0.76 | 0.17-3.47 | 0.727 |
| Prior antiplatelet treatment | 0.55 | 0.13-2.40 | 0.429 |
| Prior antihypertensive treatment | 0.51 | 0.11-2.45 | 0.400 |
| mTICI score | 2.14 | 0.85-5.39 | 0.106 |
| **Model 4** |  |  |  |
| Q4 | 0.13 | 0.03-0.61 | **0.010** |
| Atrial fibrillation | 0.24 | 0.05-1.22 | 0.085 |
| Prior statin treatment | 0.76 | 0.17-3.43 | 0.722 |
| Prior antiplatelet treatment | 0.55 | 0.13-2.38 | 0.423 |
| Prior antihypertensive treatment | 0.49 | 0.10-2.39 | 0.378 |
| Stroke onset-to-groin puncture time | 1.00 | 1.00-1.01 | 0.742 |
| mTICI score | 2.21 | 0.84-5.38 | 0.114 |

aOR=adjusted odds ratio; ASPECTS=Alberta Stroke Program Early CT Score; CI=confidence interval; mTICI=modified Thrombolysis in Cerebral Infarction: Q4=highest cEPC count quartile.

## Discussion

In patients with LVO-AIS who received EVT, we found that post-EVT cEPC mobilization was not associated with functional outcomes at three months’ follow-up. We also found that a higher cEPC count in the first 24 hours after stroke onset was associated with poor baseline CC. Our study provides new evidence regarding the prognostic role of cEPCs in the AIS setting in several ways.

First, our findings – pointing to a non-association between cEPC count in the acute LVO-AIS phase (cEPC counts on days 1 and 7, and the difference between them) and functional outcomes after three months – conflict with previous studies that report an association between cEPC count on day 7 post-stroke and better functional outcomes[10–12,18,20]. However, our study is not directly comparable to those previous studies, as discrepancies are possibly due to study population differences. Prior studies focused on minor stroke in patients who did not receive EVT[11,18,20], while our study included the severe AIS secondary to LVO in patients who received EVT. Patient recovery is also influenced by factors such as age, neurological deficit as measured by the NIHSS, success of recanalization, time from onset to treatment, and peri- and post-procedure complications[21]. Furthermore, the association reported in some studies between functional outcomes and cEPC counts was only found for specific stroke etiologies[18,20,22]. Thus, although it is well known that the cEPC count increases after an ischemic stimulus[1], the effect may not be strong enough to independently predict the functional prognosis of patients with LVO-AIS.

Second, we observed that a higher cEPC count within 24 hours in patients who received EVT was associated with poorer CC in the baseline CTA; this may be explained by the fact that we analysed early-outgrowth cEPCs, which are closely linked to the ischemic stimulus, rather than late-outgrowth cEPCs. Early-outgrowth cEPCs participate in endothelial cell regeneration by releasing growth factors, whereas late-outgrowth cEPCs differentiate into mature endothelial cells that contribute to de-novo blood vessel formation[23,24]. To detect those cEPC types, we used the CD34, VEGFR2, and CD133 surface markers, the last of which is a specific marker for early-outgrowth EPCs[25]. In physiological conditions, cEPC concentrations in blood are very low (0.0001-0.01% of mononuclear cells) to the point that they are practically undetectable[26]. However, early mobilization occurs after an ischemic stimulus. In LVO-AIS, CC is crucial to cerebral blood flow to the ischemic area[16] in the first hours post-stroke, with poor CC linked to a more severe course and poorer outcomes[27,28]. The association we found may be explained by a greater ischemic stimulus through fast infarct progression, due precisely to poor CC. Severe hypoxic conditions activate hypoxia-inducible factor-1 (HIF-1), which induces early cEPC mobilization to promote new vessel formation, and a massive release of angiogenic and anti-inflammatory factors to maintain the viability of the ischemic area[29]; additionally, angiogenic factors and inflammatory cytokines accumulated in damaged tissue attract more cEPCs[29]. cEPC concentrations are consequently elevated in more severely affected patients. It is also well known that cEPCs have vasodilatory[14] and anti-inflammatory[15] properties. However, our results would suggest that a high cEPC count in the 24 hours post-stroke does not seem to be associated with better CC, and consequently, with a better functional outcome, but rather, that this early mobilization probably reflects severe ischemia.

Third, we found new variables associated with cEPC mobilization. Patients with atrial fibrillation experienced less cEPCs mobilization at 24 hours and on day 7 after stroke onset. Supporting our findings are Siu et al. [30], who reported an association between persistent atrial fibrillation and lower cEPC counts. While it is well known that, due to the resulting proinflammatory environment, patients with atrial fibrillation are more susceptible to endothelial dysfunction[31], little is known about the atrial fibrillation link with cEPCs; one hypothesis is that cEPCs may facilitate the restoration of endothelial integrity by replacing damaged cells, leading to a reduction in cEPC levels[32]. We also found that previous treatment with anticoagulants and hospital admission in the first 6 hours after stroke onset are associated with lower cEPCs mobilization on day 7. Although the relationship between anticoagulants and cEPCs is not fully understood, Papadaki et al. [33] reported that factor Xa and thrombin activate late-outgrowth cEPCs. Anticoagulants that reduce late-outgrowth cEPCs by inhibiting thrombin and factor Xa may also reduce early-outgrowth cEPCs through the same mechanism. Since patients with atrial fibrillation usually receive anticoagulant treatment, the observed association between anticoagulant treatment and cEPCs could be also a consequence of atrial fibrillation. As for the association between cEPC count and stroke onset-to-door time <6 hours, a possible explanation could be earlier treatment that, in reducing ischemic damage, in turn reduces cEPC mobilization. Regarding intravenous fibrinolysis therapy, this was associated with greater cEPC mobilization on day 7. Supporting this finding are Sobrino et al. [34], who reported that patients treated with citicoline combined with intravenous fibrinolysis had higher cEPC counts compared with patients treated with citicoline alone. Intravenous fibrinolysis, associated with matrix metalloproteinase-9 expression[35], has been reported to be a stimulator of cEPC mobilization[36]. However, further studies of this mechanism are needed.

In view of our results, we are of the opinion that cEPC count in LVO-AIS after EVT may not serve as a reliable biomarker to predict functional outcomes. Clearly, other prominent clinical variables, such as baseline NIHSS and baseline ASPECTS scores, outweigh the potential effect of cEPCs. Additionally, as discussed above, we believe that the association between cEPCs and CC reflects a greater ischemic stimulus rather than any direct relationship between cEPCs and vessels participating in CC. Therefore, we would argue against the use of early cEPC counts to predict clinical outcomes in this population in future studies.

## Limitations

Some limitations of our study are that it was a single-centre study and was based on a relatively small sample size, especially of patients analysed on day 7. In addition, some patients were lost to follow-up, there are missing data for some variables, and CC was measured at baseline whereas cEPC count was measured at 24±12 hours and on day 7±1 from stroke onset. Finally, data on infarct volume were not available to confirm our hypothesis regarding the association between cEPCs and LVO-AIS functional outcomes.

## Conclusions

In patients with AIS secondary to anterior circulation LVO who received EVT, we found no association between cEPC mobilization and functional outcomes, but found that early cEPC mobilization (within 24 hours) was associated with poor baseline CC.

## ABBREVIATIONS AND ACRONYMS

AIS: acute ischemic stroke
ASPECTS: Alberta Stroke Program Early CT score
cEPC: circulating endothelial progenitor cell
CC: collateral circulation
CS: collateral score
CT: computed tomography
CTA: computed tomography angiography
EDTA: ethylene diamine tetra acetic acid
EVT: endovascular therapy
HIF-I: hypoxia-inducible factor-1
LVO: large-vessel occlusion
MACS: magnetic-activated cell sorting
MCA: middle cerebral artery
mRS: modified Rankin Scale
mTICI: modified Thrombolysis in Cerebral Infarction Scale
NIHSS: National Institutes of Health Stroke Scale
TICA: terminal internal carotid artery

## Acknowledgments

We are grateful to citometry platform of IR SANT PAU for the assistance with the cytometry analysis, to Ailish Maher for the help with language, and to Biochemistry and Molecular Biology department of Universitat Autònoma de Barcelona for their support in this research.

## Statement of Ethics

The study protocol was approved by the Ethics Committee oh Hospital de la Santa Creu i Sant Pau, Barcelona, Spain (reference number: IIBSP-COL-2019-64). The study was conducted in accordance with the principles of the Declaration of Helsinki. Written informed consent to participate was obtained from all patients or, when this was not possible, from their legally authorized representatives. No potentially identifiable images or other personal data are included in this article.

## Author Contributions

All authors have seen and approved the manuscript.

Conceptualization: J.M.-F., P.C.-R.

Methodology: A.A.-S., P.C.-R., N.P., E.J.-X.

Resources: A.A.-S., P.C.-R., M.G.-J., N.P., E.J.-X., R.M., C.G.-F., G.E.-D., A.L.-G., A.M.-D., L.P.-S., A.R.-P., J.M.-F.

Project administration: A.A.-S., P.C.-R., M.G.-J., C.G.-F., G.E.-D., A.L.-G., A.M.-D., L.P.-S., A.R.-P., J.M.-F.

Formal analysis: A.A.-S., P.C.-R

Investigation: A.A.-S., P.C.-R., M.G.-J., N.P., E.J.-X., R.M., M.S., J.P.M.-G., J.O.-Q., J.M.-F.

Data curation: A.A.-S., P.C.-R

Writing – original draft: A.A.-S., P.C.-R.

Writing – review and editing: all authors.

Supervision: J.M.-F., P.C.-R.

Funding acquisition: J.M.-F.

## Data Availability Statement

All data generated or analysed during this study are included in this article and its supplementary material files. Further enquiries can be directed to the corresponding author.

