## Supplemental Table 1 for "Association of circulating endothelial progenitor cell count and functional outcome in patients with acute ischemic stroke due to intracranial large vessel occlusion"

**Supplementary Table S1 Bivariate analysis of cEPC predictors according to cEPC presence or absence at 24 hours**

|  | **cEPCs- 24h (n=48)** | **cEPCs+ 24h (n=42)** | **p** |
| --- | --- | --- | --- |
| Age, mean (SD) | 71.8 (13.5) | 74.7 (11.6) | 0.405 |
| Sex (women), n (%) | 20 (41.7) | 17 (40.5) | 0.909 |
| Current smokers, n (%) | 9 (18.8) | 8 (19.1) | 0.971 |
| Obesity (BMI ≥30), n (%) | 10 (20.8) | 6 (14.3) | 0.418 |
| Hypertension, n (%) | 29 (60.4) | 33 (78.6) | 0.063 |
| Diabetes, n (%) | 12 (25.0) | 9 (21.4) | 0.689 |
| Dyslipidemia, n (%) | 23 (47.9) | 25 (59.5) | 0.271 |
| Previous stroke, n (%) | 9 (18.8) | 7 (16.7) | 0.796 |
| Heart failure, n (%) | 4 (8.3) | 6 (14.3) | 0.370 |
| Atrial fibrillation, n (%) | 17 (35.4) | 7 (16.7) | **0.045** |
| Prior antiplatelet treatment, n (%) | 13 (27.1) | 11 (26.2) | 0.924 |
| Prior statin treatment, n (%) | 23 (47.9) | 16 (38.1) | 0.348 |
| Prior anticoagulant treatment, n (%) | 14 (29.2) | 10 (23.8) | 0.566 |
| Prior antihypertensive treatment, n (%) | 30 (62.5) | 29 (69.1) | 0.514 |
| Previous mRS score, median (IQR) | 0 (0-2) | 0 (0-1) | 0.818 |
| Baseline NIHSS, median (IQR) | 17 (11-20) | 18 (11-21) | 0.585 |
| ASPECTS, median (IQR)* | 9 (8-10) | 8 (7-10) | 0.257 |
| Stroke onset-to-door <6 hours, n (%) | 36 (75.0) | 28 (66.7) | 0.384 |
| Stroke onset-to-groin puncture (minutes), median (IQR) | 65 (54-74) | 66 (55-87) | 0.555 |
| Occlusion, n (%) |  |  |  |
| M1 | 24 (50.0) | 24 (57.1) | 0.792 |
| M2 | 15 (31.28) | 11 (26.2) |  |
| TICA | 9 (18.8) | 7 (16.7) |  |
| Stroke etiology, n (%)* |  |  |  |
| Large-artery atherosclerosis | 7 (14.6) | 9 (22.0) | 0.649 |
| Cardioembolism | 23 (47.9) | 18 (43.9) |  |
| Uncommon | 5 (10.4) | 2 (4.9) |  |
| Undetermined | 13 (27.1) | 12 (29.3) |  |
| Reperfusion treatment, n (%) |  |  |  |
| Mechanical thrombectomy | 28 (58.3) | 31 (73.8) | 0.123 |
| Intravenous fibrinolysis + mechanical thrombectomy | 20 (41.7) | 11 (26.2) |  |
| mTICI score ≥2b, n (%) ** | 44 (93.6) | 41 (100.0) | 0.100 |
| Good CC, n (%)*** | 37 (78.7) | 28 (73.7) | 0.586 |

*, **, ***: data available for 89, 88, and 85 patients, respectively. Good CC=collateral score 2-3. ASPECTS=Alberta Stroke Program Early CT Score; BMI=body mass index; CC=collateral circulation; cEPC=circulating endothelial progenitor cell; cEPC- 24h=absence of cEPCs at 24 hours; cEPCs+ 24h=presence of cEPCs at 24 hours; IQR=interquartile range; mRS=modified Rankin Scale; mTICI=modified Thrombolysis in Cerebral Infarction; NIHSS=National Institute of Health Stroke Scale; TICA=terminal internal carotid artery; SD=standard deviation.
