## Supplemental Table 2 for "Association of circulating endothelial progenitor cell count and functional outcome in patients with acute ischemic stroke due to intracranial large vessel occlusion"

**Supplementary Table S2 Bivariate analysis of cEPC predictors according to cEPC presence or absence on day 7**

|  | **cEPCs- d7 (n=35)** | **cEPCs+ d7 (n=27)** | **p** |
| --- | --- | --- | --- |
| Age, mean (SD) | 72.3 (11.8) | 71.6 (15.8) | 0.966 |
| Sex (women), n (%) | 11 (31.4) | 14 (51.9) | 0.104 |
| Current smokers, n (%) | 5 (14.3) | 7 (25.9) | 0.250 |
| Obesity (BMI ≥30), n (%) | 7 (20.0) | 6 (22.2) | 0.831 |
| Hypertension, n (%) | 23 (65.7) | 20 (74.1) | 0.479 |
| Diabetes, n (%) | 12 (34.3) | 5 (18.5) | 0.168 |
| Dyslipidemia, n (%) | 20 (57.1) | 12 (44.4) | 0.321 |
| Previous stroke, n (%) | 7 (20.0) | 5 (18.5) | 0.884 |
| Heart failure, n (%) | 3 (8.6) | 3 (11.1) | 0.737 |
| Atrial fibrillation, n (%) | 12 (34.3) | 3 (11.1) | **0.035** |
| Prior antiplatelet treatment, n (%) | 10 (28.6) | 7 (25.9) | 0.817 |
| Prior statin treatment, n (%) | 18 (51.4) | 8 (29.6) | 0.085 |
| Prior anticoagulant treatment, n (%) | 13 (37.1) | 2 (7.4) | **0.007** |
| Prior antihypertensive treatment, n (%) | 23 (65.7) | 18 (66.7) | 0.937 |
| Previous mRS score, median (IQR) | 0 (0-1) | 0 (0-1) | 0.387 |
| Baseline NIHSS, median (IQR) | 17 (11-21) | 17 (8-21) | 0.560 |
| ASPECTS, median (IQR) | 9 (8-10) | 8 (7-10) | 0.324 |
| Stroke onset-to-door <6 hours, n (%) | 17 (48.6) | 5 (18.5) | **0.014** |
| Stroke onset-to-groin puncture (minutes), median (IQR) | 67 (55-78) | 66 (55-89) | 0.977 |
| Occlusion, n (%) |  |  |  |
| M1 | 16 (45.7) | 14 (51.9) | 0.724 |
| M2 | 11 (31.4) | 9 (33.3) |  |
| TICA | 8 (22.9) | 4 (14.8) |  |
| Stroke etiology, n (%) |  |  |  |
| Large-artery atherosclerosis | 5 (14.3) | 7 (25.9) | 0.090 |
| Cardioembolism | 19 (54.3) | 7 (25.9) |  |
| Uncommon | 4 (11.4) | 2 (7.41) |  |
| Undetermined | 7 (20.0) | 11 (40.7) |  |
| Reperfusion therapy, n (%) |  |  |  |
| Mechanical thrombectomy | 28 (80.0) | 12 (44.4) | **0.004** |
| Intravenous fibrinolysis + mechanical thrombectomy | 7 (20.0) | 15 (55.6) |  |
| mTICI score ≥2b, n (%)* | 34 (100.0) | 25 (95.2) | 0.249 |
| Good CC, n (%)** | 28 (82.4) | 17 (70.8) | 0.300 |

*, **: data available for 60 and 58 patients, respectively. Good CC=collateral score 2-3. ASPECTS=Alberta Stroke Program Early CT Score; BMI=body mass index; CC=collateral circulation; cEPC=circulating endothelial progenitor cell; cEPCs+ d7=presence of cEPCs on day 7; cEPCs- d7=absence of cEPCs on day 7; IQR=interquartile range; mRS=modified Rankin Scale; mTICI=modified Thrombolysis in Cerebral Infarction; NIHSS=National Institute of Health Stroke Scale; SD=standard deviation; TICA=terminal internal carotid artery.
