## Supplemental Table 3 for "Association of circulating endothelial progenitor cell count and functional outcome in patients with acute ischemic stroke due to intracranial large vessel occlusion"

**Supplementary Table S3 Bivariate analysis of cEPC predictors according to cEPC increase or decrease in the first week**

|  | **ΔcEPCs- (n=47)** | **ΔcEPCs+ (n=15)** | **p** |
| --- | --- | --- | --- |
| Age, mean (SD) | 71.9 (11.5) | 72.5 (19.2) | 0.411 |
| Sex (women), n (%) | 16 (34.0) | 9 (60.0) | 0.074 |
| Current smoking, n (%) | 10 (21.3) | 2 (13.3) | 0.498 |
| Obesity (BMI ≥30), n (%) | 9 (19.2) | 4 (26.7) | 0.533 |
| Hypertension, n (%) | 32 (68.1) | 11 (73.3) | 0.701 |
| Diabetes, n (%) | 13 (27.7) | 4 (26.7) | 0.940 |
| Dyslipidaemia, n (%) | 26 (55.3) | 6 (40.0) | 0.301 |
| Previous stroke, n (%) | 9 (19.2) | 3 (20.0) | 0.942 |
| Heart failure, n (%) | 4 (8.5) | 2 (13.3) | 0.582 |
| Atrial fibrillation, n (%) | 12 (25.5) | 3 (20.0) | 0.663 |
| Prior antiplatelet treatment, n (%) | 12 (25.5) | 5 (33.3) | 0.555 |
| Prior statin treatment, n (%) | 23 (48.9) | 3 (20.0) | 0.071 |
| Prior anticoagulant treatment, n (%) | 13 (27.7) | 2 (13.3) | 0.259 |
| Prior antihypertensive treatment, n (%) | 31 (66.0) | 10 (66.7) | 0.960 |
| Previous mRS score, median (IQR) | 0 (0-1) | 0 (0-1) | 0.932 |
| Baseline NIHSS, median (IQR) | 17 (11-21) | 13 (5-21) | 0.273 |
| ASPECTS, median (IQR) | 8 (7-10) | 9 (8-10) | 0.925 |
| Stroke onset-to-door <6 hours, n (%) | 21 (44.7) | 1 (6.7) | **0.007** |
| Stroke onset-to-groin puncture (minutes), median (IQR) | 67 (56-83) | 60 (53-72) | 0.406 |
| Occlusion, n (%) |  |  |  |
| M1 | 22 (46.8) | 8 (53.3) | 0.787 |
| M2 | 15 (31.9) | 5 (33.3) |  |
| TICA | 10 (21.3) | 2 (13.3) |  |
| Stroke etiology, n (%) |  |  |  |
| Large-artery atherosclerosis | 8 (17.0) | 4 (26.7) | 0.128 |
| Cardioembolism | 22 (46.8) | 4 (26.7) |  |
| Uncommon | 6 (12.8) | 0 (0.0) |  |
| Undetermined | 11 (23.4) | 7 (46.7) |  |
| Reperfusion therapy, n (%) |  |  |  |
| Mechanical thrombectomy | 35 (74.5) | 5 (33.3) | **0.004** |
| Intravenous fibrinolysis + mechanical thrombectomy | 12 (25.5) | 10 (66.7) |  |
| mTICI score ≥2b, n (%)* | 46 (100.0) | 13 (92.9) | 0.068 |
| Good CC, n (%)** | 35 (77.8) | 10 (76.9) | 0.948 |

*, **: data available for 60 and 58 patients, respectively. Good CC=collateral score 2-3. ΔcEPC= change in cEPCs from 24 hours to day 7 (+ increase; - decrease); ASPECTS=Alberta Stroke Program Early CT Score; BMI=body mass index; CC=collateral circulation; cEPC=circulating endothelial progenitor cell; IQR=interquartile range; mRS=modified Rankin Scale; mTICI=modified Thrombolysis in Cerebral Infarction; NIHSS=National Institute of Health Stroke Scale; SD=standard deviation; TICA=terminal internal carotid artery.
