## Supplemental Table 4 for "Association of circulating endothelial progenitor cell count and functional outcome in patients with acute ischemic stroke due to intracranial large vessel occlusion"

**Supplementary Table S4 Bivariate stratified analysis of cEPC predictors according to the highest cEPC count <24 hours (Q4)**

|  | **Q1-Q3 (n=44)** | **Q4 (n=13)** | **p** |
| --- | --- | --- | --- |
| Age, mean (SD) | 72.4 (13.0) | 69.2 (10.4) | 0.238 |
| Sex (women), n (%) | 14 (31.8) | 6 (46.2) | 0.341 |
| Current smoking, n (%) | 9 (20.5) | 3 (23.1) | 0.839 |
| Obesity (BMI≥30), n (%) | 10 (22.7) | 1 (7.7) | 0.227 |
| Hypertension, n (%) | 29 (65.9) | 8 (61.5) | 0.772 |
| Diabetes, n (%) | 9 (20.5) | 3 (23.1) | 0.807 |
| Dyslipidaemia, n (%) | 22 (50.0) | 9 (69.2) | 0.221 |
| Previous stroke, n (%) | 8 (18.2) | 1 (7.7) | 0.668 |
| Heart failure, n (%) | 9 (20.5) | 0 (0.0) | 0.101 |
| Atrial fibrillation, n (%) | 12 (27.3) | 3 (23.1) | 0.763 |
| Prior antiplatelet treatment, n (%) | 15 (34.1) | 2 (15.4) | 0.304 |
| Prior statin treatment, n (%) | 22 (50.0) | 5 (38.5) | 0.464 |
| Prior anticoagulant treatment, n (%) | 11 (25.0) | 3 (23.1) | 0.887 |
| Prior antihypertensive treatment, n (%) | 31 (70.5) | 6 (46.2) | 0.107 |
| Previous mRS score, median (IQR) | 0 (0-2) | 0 (0-2) | 0.835 |
| Baseline NIHSS, median (IQR) | 19 (12-22) | 18 (13-21) | 0.856 |
| ASPECTS, median (IQR)* | 9 (8-10) | 9 (7-10) | 0.414 |
| Stroke onset-to-door <6 hours, n (%) | 38 (86.4) | 12 (92.3) | 0.566 |
| Stroke onset-to-groin puncture (minutes), median (IQR) | 127 (104-223) | 160 (98-274) | 0.955 |
| Occlusion, n (%) |  |  |  |
| M1 | 25 (56.8) | 6 (46.15) | 0.760 |
| M2 | 12 (27.3) | 5 (38.5) |  |
| TICA | 7 (15.9) | 2 (15.4) |  |
| Stroke etiology, n (%)* |  |  |  |
| Large-artery atherosclerosis | 7 (16.3) | 3 (23.1) | 0.938 |
| Cardioembolism | 20 (46.5) | 5 (38.5) |  |
| Uncommon | 3 (7.0) | 1 (7.7) |  |
| Undetermined | 13 (30.2) | 4 (30.8) |  |
| Reperfusion therapy, n (%) |  |  |  |
| Mechanical thrombectomy | 23 (52.3) | 8 (61.5) | 0.556 |
| Intravenous fibrinolysis + mechanical thrombectomy | 21 (47.7) | 5 (38.5) |  |
| mTICI score ≥2b, n (%) | 43 (97.7) | 13 (100.0) | 0.583 |
| Good CC, n (%) | 34 (77.3) | 6 (46.2) | **0.031** |

*: data available for 56 patients. Q1-Q3= cEPC count quartiles 1 to 3; Q4=highest cEPC count quartile. Good CC=collateral score 2-3. ASPECTS=Alberta Stroke Program Early CT Score; BMI=body mass index; CC=collateral circulation; cEPC=circulating endothelial progenitor cell; IQR=interquartile range; mRS=modified Rankin Scale; mTICI=modified Thrombolysis in Cerebral Infarction; NIHSS=National Institute of Health Stroke Scale; SD=standard deviation; TICA=terminal internal carotid artery.
