## Supplemental Table 5 for "Association of circulating endothelial progenitor cell count and functional outcome in patients with acute ischemic stroke due to intracranial large vessel occlusion"

**Supplementary Table S5 Bivariate analysis of predictors of poor or good CC**

|  | **Poor CC**  **(n=17)** | **Good CC (n=40)** | **p** |
| --- | --- | --- | --- |
| Age, mean (SD) | 70.4 (11.53) | 72.2 (13.0) | 0.535 |
| Sex (women), n (%) | 6 (35.3) | 14 (35.0) | 0.983 |
| Current smoking, n (%) | 4 (23.5) | 8 (20.0) | 0.765 |
| Obesity (BMI≥30), n (%) | 4 (23.5) | 7 (17.5) | 0.598 |
| Hypertension, n (%) | 12 (70.6) | 25 (62.5) | 0.558 |
| Diabetes, n (%) | 4 (23.5) | 8 (20.0) | 0.300 |
| Dyslipidemia, n (%) | 9 (52.9) | 22 (55.0) | 0.886 |
| Previous stroke, n (%) | 2 (11.8) | 7 (17.5) | 0.710 |
| Heart failure, n (%) | 4 (23.5) | 5 (12.5) | 0.428 |
| Atrial fibrillation, n (%) | 7 (41.2) | 8 (20.0) | 0.097 |
| Prior antiplatelet treatment, n (%) | 6 (35.3) | 11 (27.5) | 0.547 |
| Prior statin treatment, n (%) | 10 (58.8) | 17 (42.5) | 0.259 |
| Prior anticoagulant treatment, n (%) | 6 (35.3) | 8 (20.0) | 0.220 |
| Prior antihypertensive treatment, n (%) | 12 (70.6) | 25 (62.5) | 0.558 |
| Previous mRS score, median (IQR) | 0 (0-1) | 0 (0-2) | 0.999 |
| Baseline NIHSS, median (IQR) | 19 (11-23) | 18 (13-21) | 0.701 |
| ASPECTS, median (IQR) | 8 (7-10) | 9 (8-10) | 0.068 |
| Stroke onset-to-door <6 hours, n (%) | 14 (82.4) | 36 (90.0) | 0.421 |
| Stroke onset-to-groin puncture (minutes), median (IQR) | 160 (109-202) | 121 (99-234) | 0.663 |
| Occlusion, n (%) |  |  |  |
| M1 | 12 (70.6) | 19 (47.5) | 0.248 |
| M2 | 4 (23.5) | 13 (32.5) |  |
| TICA | 1 (5.9) | 8 (20.0) |  |
| Stroke etiology, n (%) * |  |  |  |
| Large-artery atherosclerosis | 2 (11.8) | 8 (20.5) | 0.864 |
| Cardioembolism | 9 (52.9) | 16 (41.0) |  |
| Uncommon | 1 (5.9) | 3 (7.7) |  |
| Undetermined | 5 (29.4) | 12 (30.8) |  |
| Reperfusion therapy, n (%) |  |  |  |
| Mechanical thrombectomy | 12 (70.6) | 19 (47.5) | 0.109 |
| Intravenous fibrinolysis + mechanical thrombectomy | 5 (29.4) | 21 (52.5) |  |
| mTICI score ≥2b, n (%) | 17 (100.0) | 39 (97.5) | 0.511 |

*: data available for 56 patients. ASPECTS=Alberta Stroke Program Early CT Score; BMI=body mass index; CC=collateral circulation; IQR=interquartile range; mRS=modified Rankin Scale; mTICI=modified thrombolysis in cerebral infarction; NIHSS=National Institute of Health Stroke Scale; SD=standard deviation; TICA=terminal internal carotid artery.
